# Within-Year PM_2.5_ Exposure Structure Provides Mortality-Predictive Information Beyond the Annual Mean

**DOI:** 10.64898/2026.09.20.26363510

**Authors:** Ren Zhang

## Abstract

Long-term PM_2.5_ exposure is associated with increased mortality and is usually represented by annual mean concentration, which does not capture how concentrations are distributed across days. We tested whether the prespecified PM_2.5_ magnitude-rank index (PMRI), a summary of within-year daily concentration structure, improves mortality prediction beyond the annual mean. Daily modeled PM_2.5_ estimates were linked to age-adjusted mortality rates for 27,289 county-years from 3,064 U.S. counties during 2003–2011. PMRI is the largest *k* for which at least *k* days reached *k* µg/m³. County-grouped 10-fold cross-validation compared models with annual mean alone versus annual mean plus PMRI, with additional geographic, temporal, cause-specific, and specification analyses. Annual mean PM_2.5_ had the highest standalone predictive performance. Adding PMRI increased county-held-out R² from 0.0802 to 0.1391 (ΔR² = 0.0589; 95% CI, 0.0499–0.0677) and reduced RMSE and MAE. Improvement persisted across geographic and temporal validations and all six cause-specific mortality outcomes. Within-year PM_2.5_ exposure structure therefore contains reproducible mortality-predictive information not captured by annual mean concentration.

## 1. Introduction

Long-term exposure to fine particulate matter (PM_2.5_) is consistently associated with increased mortality. Prospective cohorts and large population-based analyses have reported associations with all-cause and cause-specific mortality, including at relatively low concentrations (Dockery, Pope et al. 1993, Pope Iii, Burnett et al. 2002, Crouse, Peters et al. 2012, Di, Wang et al. 2017). Systematic reviews and meta-analyses, including the recent update prepared for the World Health Organization Global Air Quality Guidelines, confirm a large and expanding evidence base (Chen and Hoek 2020, Orellano, Kasdagli et al. 2024). Accordingly, annual or multiyear mean concentration is the conventional representation of long-term PM_2.5_ exposure in epidemiologic studies.

Annual mean PM_2.5_ captures overall exposure level but not its distribution across days. The same annual mean may reflect persistent moderate concentrations or lower background concentrations punctuated by brief high-concentration episodes; these profiles differ in the magnitude and frequency of elevated daily concentrations. This distinction is reflected in World Health Organization air quality guidelines and U.S. national standards, both of which include annual and 24-hour PM_2.5_ metrics (Bachmann, Damberg et al. 1996, WHO 2021). Mortality associations have also been observed over both short-and long-term exposure periods, including when acute and chronic effects were across (Shi, Zanobetti et al. 2016, Liu, Chen et al. 2019). Previous studies have therefore examined PM_2.5_ characteristics not represented by the mean. In a time-series analysis, the standard deviation of daily PM_2.5_ was associated with respiratory mortality after adjustment for daily mean concentration (Lin, Ma et al. 2016). A cardiovascular cohort study that included both annual mean PM_2.5_ and the annual frequency of high-concentration exposure reported associations for both metrics (Cho, Kang et al. 2022). Wildfire studies have characterized long-term exposure using mean concentration, peak intensity, exposed days or weeks, and smoke-wave occurrence (Casey, Kioumourtzoglou et al. 2024, Schwarz, Frankland et al. 2025), while wildfire event days may modify the association between long-term all-source PM_2.5_ and mortality (Spoto, Dominici et al. 2026). Recent analyses at shorter temporal scales likewise suggest that exposure variance and extreme pollution days contain mortality-relevant information not captured by average concentration alone (Deng, Zhou et al. 2026, Ghaddar, Babiarz et al. 2026). However, it remains to be established whether a daily-derived annual summary of all-source PM_2.5_ improves mortality prediction beyond the annual mean and whether any such improvement generalizes across locations and time.

To address this gap, we used the recently developed PM_2.5_ magnitude-rank index (PMRI), a prespecified summary of the magnitude-frequency structure of ranked daily concentrations (Zhang 2026). PMRI was defined independently of health outcomes and was not optimized for the present analysis. In a nationwide county-year study, we assessed whether adding PMRI to annual mean PM_2.5_ improved prediction of age-adjusted all-cause mortality in held-out counties. We further evaluated generalization across geographic partitions, time periods, and causes of death and assessed robustness to flexible modeling of the annual mean and inclusion of the annual 98th percentile.

## 2. Materials and Methods

### 2.1 Study design and data sources

We conducted a nationwide longitudinal ecological study of U.S. counties from 2003 through 2011, with county-year as the unit of analysis. Daily county-level PM_2.5_ estimates were summarized annually and linked to county-year age-adjusted mortality rates by five-digit county Federal Information Processing Standards code and calendar year. The analysis was designed to quantify the incremental out-of-sample predictive performance of alternative annual PM_2.5_ summaries and was not intended to estimate individual-level or causal effects.

Daily county-level PM_2.5_ estimates were obtained from the National Environmental Public Health Tracking Network of the Centers for Disease Control and Prevention (CDC) (CDC). The estimates were generated with the U.S. Environmental Protection Agency Downscaler, which combines air-quality monitoring observations with output from the Community Multiscale Air Quality model (Berrocal, Gelfand et al. 2010). The primary exposure variable, PM25_POP_PRED, was the population-weighted county mean of predicted 24-hour PM_2.5_ concentration, expressed in µg/m³. Each record was identified by five-digit county FIPS code and calendar date. The source data comprised 10,219,283 county-day observations during 2003–2011 before linkage to mortality outcomes.

County-year all-cause and cause-specific mortality data were obtained from the CDC WONDER Compressed Mortality File (National_Center_for_Health_Statistics). The primary outcome was all-cause mortality. Cause-specific outcomes were cardiovascular or circulatory mortality (ICD-10 I00–I99), ischemic heart disease (I20–I25), acute myocardial infarction (I21–I22), stroke or cerebrovascular disease (I60–I69), respiratory mortality (J00–J99), and chronic lower respiratory disease (J40–J47). For each outcome, we extracted the reported age-adjusted mortality rate per 100,000 population. Suppressed, missing, or unreliable rates were excluded without imputation. Because data availability and reliability varied across outcomes, each cause-specific analysis used its own eligible cohort rather than a common cohort restricted to observations available for all outcomes. After exposure–mortality linkage and these exclusions, the primary all-cause cohort comprised 27,289 county-years.

### 2.2 Exposure completeness and annual metrics

A county-year was eligible if valid estimates were available for at least 90% of the 365 expected days (366 in leap years). Annual summaries were calculated from all available days in eligible county-years; missing daily values were not imputed. County-year identifiers were required to be unique. These criteria yielded 27,981 county-years before application of mortality reliability criteria.

Annual mean PM_2.5_ was the benchmark exposure metric. For PMRI, daily concentrations within each county-year were ordered from highest to lowest, x₍₁₎ ≥ x₍₂₎ ≥…≥ x₍ₙ₎, and the integer index was defined as [18]:

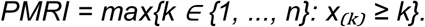

Thus, PMRI equals k when at least k days have PM_2.5_ concentrations of k µg/m³ or greater. The published integer PMRI remained the primary measure. For sensitivity analysis, a continuous PMRI was defined as the fractional rank at which the linearly interpolated ranked concentration curve crossed the identity line between the two adjacent ranks surrounding the integer PMRI. The metric was defined independently of health outcomes and was not optimized in the present study. Conventional comparators were the annual 98th percentile (P98), annual maximum, and numbers of days with concentrations of at least 15 or 35 µg/m³.

### 2.3 Prediction models and performance measures

We compared two ordinary least-squares prediction models. The base model included annual mean PM_2.5_ alone, whereas the augmented model included annual mean PM_2.5_ and PMRI:

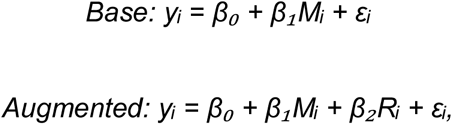

where yᵢ is the age-adjusted mortality rate for county-year i, Mᵢ is annual mean PM_2.5_, and Rᵢ is PMRI. The primary estimand was the increment in pooled out-of-sample coefficient of determination, ΔR² = R²augmented − R²base. For each validation analysis, R² was calculated from the concatenated held-out predictions as 1 minus the residual sum of squares divided by the total sum of squares. Root mean squared error (RMSE) and mean absolute error (MAE) were secondary performance measures; negative augmented-minus-base differences in RMSE or MAE favored the augmented model. Incremental prediction was evaluated from held-out performance. Because these exposure-only models did not estimate a confounder-adjusted association, their coefficients were not interpreted etiologically.

To assess robustness to specification of the annual-mean term, we repeated the comparison using quadratic and cubic polynomial functions and a three-knot cubic spline. Each specification was fitted within the training data and applied unchanged to the corresponding held-out observations. We also evaluated whether PMRI improved prediction after inclusion of annual mean and P98.

### 2.4 Geographic validation and uncertainty

County-grouped 10-fold cross-validation was the primary validation design. All years from a county were assigned to the same fixed fold, so no county contributed observations to both the training and test data within a fold. In each iteration, the two models were fitted in nine folds and evaluated in the omitted fold. Held-out predictions were then pooled across all 10 folds for calculation of R², RMSE, and MAE.

We evaluated geographic transportability in two additional designs. In state-grouped 10-fold cross-validation, each state and all of its county-years were retained in a single fixed fold. In leave-one-state-out validation, the models were trained on all other states and evaluated in the omitted state. States were eligible for the latter analysis if they had at least 30 observations and nonzero outcome variance; 47 states met these criteria. The primary leave-one-state-out estimate was calculated from all pooled held-out predictions, while state-specific performance was retained to characterize heterogeneity.

Confidence intervals for ΔR² were obtained by paired cluster bootstrap resampling of the held-out predictions. Counties were resampled for county-held-out analyses and states for state-held-out analyses; all county-years belonging to a sampled cluster were retained together.

Performance for the base and augmented models was recalculated within the same resample. Primary analyses used 4,000 replicates with random seed 20260828; cause-specific analyses used 5,000 replicates with archived seeds. Percentile 95% confidence intervals were defined by the 2.5th and 97.5th percentiles of the paired bootstrap distribution. The base–augmented comparison was repeated for each cause-specific outcome using county-grouped cross-validation and paired county-bootstrap intervals.

### 2.5 Temporal validation

Temporal generalization was examined by leave-one-year-out validation and by two forward-validation analyses. For leave-one-year-out validation, each calendar year from 2003 through 2011 was omitted in turn; predictions from the nine omitted years were pooled, and uncertainty was assessed by resampling years. The forward analyses trained the models on 2003–2007 and evaluated them on 2008–2011, and trained on 2003–2008 and evaluated them on 2009– 2011. In each analysis, the evaluation period followed the training period.

### 2.6 Statistical software

Analyses were conducted in Python 3.12.3 using pandas 3.0.5, NumPy 2.5.2, SciPy 1.18.1, scikit-learn 1.9.0, and statsmodels 0.15.0. Figures were generated using Matplotlib 3.10.8.

## 3. Results

### 3.1 Analytic cohort and exposure characteristics

Of the 27,981 exposure-eligible county-years, 27,289 (97.5%) from 3,064 counties had usable all-cause mortality data and comprised the primary analytic cohort. Counties contributed an average of 8.9 of the nine study years during 2003–2011 (Figure 1; Table 1). Mean age-adjusted all-cause mortality was 852.93 deaths per 100,000 population (SD, 152.02), and mean annual PM_2.5_ concentration was 10.13 µg/m³ (SD, 2.30). PMRI had a median of 18 (interquartile range, 6) and ranged from 7 to 38. Figure 1 shows broad geographic variation in mortality and annual mean PM_2.5_ across the common analytic county footprint. Among the six prespecified annual exposure summaries, annual mean PM_2.5_ was the strongest standalone predictor in county-grouped cross-validation. Its held-out R² was 0.0802, compared with 0.0446 for the number of days with PM_2.5_ ≥15 µg/m³, 0.0313 for PMRI, 0.0180 for P98, 0.0018 for annual maximum, and−0.0005 for the number of days with PM_2.5_ ≥35 µg/m³ (Table 1). Annual mean PM_2.5_ also yielded the lowest RMSE and MAE, and was therefore used as the benchmark in subsequent analyses.

**Figure 1.**
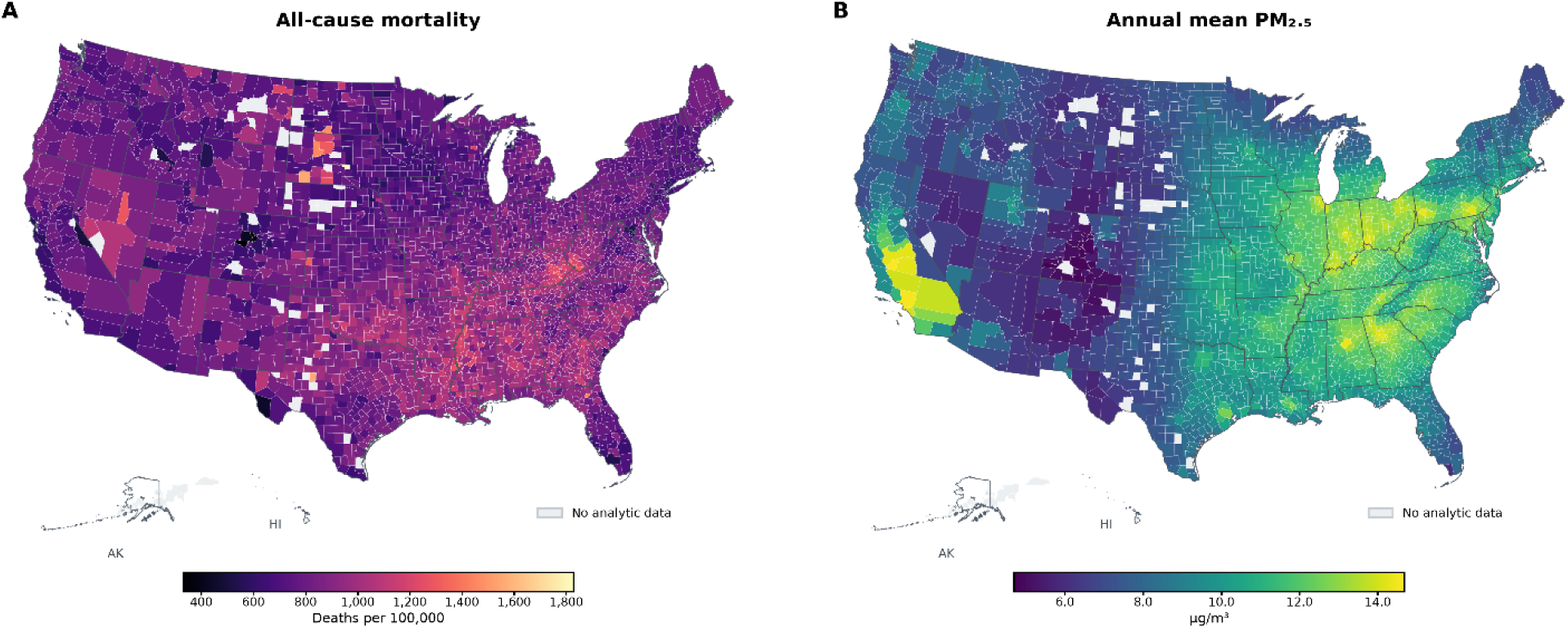
Geographic distribution of all-cause mortality and annual mean PM_2.5_ across U.S. counties. **A**, County-level age-adjusted all-cause mortality rate, calculated as the unweighted mean across included county-years during 2003–2011. **B**, County-level annual mean PM_2.5_ concentration, calculated as the unweighted mean across included county-years during the same period. Light gray indicates counties without usable analytic data. Mortality rates are expressed as deaths per 100,000 population and PM_2.5_ concentrations as µg/m³.

**Table 1.** Analytic cohort characteristics and standalone predictive performance of annual PM_2.5_ metrics.

| Variable | Mean ± SD | Median (IQR) | Range | County-held-out prediction | | |
| --- | --- | --- | --- | --- | --- | --- |
|  |  |  |  | R<sup>2</sup> | RMSE | MAE |
| All-cause mortality | 852.93 ± 152.02 | 842.20 (198.80) | 100.00–2332.70 | — | — | — |
| Annual mean PM<sub>2.5</sub> | 10.13 ± 2.30 | 10.14 (3.33) | 4.06–17.71 | 0.0802 | 145.79 | 113.48 |
| PMRI | 18.1 ± 4.2 | 18 (6) | 7–38 | 0.0313 | 149.62 | 117.14 |
| P98 | 21.99 ± 6.24 | 21.68 (8.25) | 7.73–82.26 | 0.0180 | 150.64 | 118.05 |
| Annual maximum | 30.94 ± 10.53 | 30.06 (13.13) | 10.15–221.09 | 0.0018 | 151.88 | 119.10 |
| Days $\geq 15$ $\mu\text{g}/\text{m}^3$ | 54.7 ± 40.9 | 48 (62) | 0–204 | 0.0446 | 148.59 | 116.12 |
| Days $\geq 35$ $\mu\text{g}/\text{m}^3$ | 0.94 ± 2.21 | 0 (1) | 0–46 | –0.0005 | 152.06 | 119.21 |

The primary cohort comprised 27,289 county-years from 3,064 counties during 2003–2011. County-held-out predictive performance was evaluated using county-grouped 10-fold cross-validation, with all years from a county assigned to the same validation fold. Mortality rates are age adjusted and expressed as deaths per 100,000 population; PM<sub>2.5</sub> concentrations are expressed as $\mu\text{g}/\text{m}^3$ . IQR indicates interquartile range; MAE, mean absolute error; P98, annual 98th percentile of daily PM<sub>2.5</sub>; PMRI, PM<sub>2.5</sub> magnitude-rank index; RMSE, root mean squared error; SD, standard deviation.

### 3.2 Relationship between annual mean PM_2.5_ and PMRI

Across the 27,289 county-years, PMRI was strongly correlated with annual mean PM_2.5_ (Pearson r = 0.934; Figure 2A). Nevertheless, substantial dispersion in PMRI remained among county-years with similar annual mean concentrations, indicating variation in within-year magnitude–frequency structure not represented by the mean alone. To characterize this variation without reference to mortality, same-year county-year pairs were assigned to mutually exclusive strata according to the relative difference in annual mean PM_2.5_: less than 1%, 1% to less than 5%, or 5% to 10%. Absolute PMRI differences showed upper-tailed distributions in all three strata (Figure 2B). Because individual county-years could contribute to multiple pairs, these distributions were treated descriptively, and no pair-level inferential tests were performed. Ravalli County, Montana, and Thayer County, Nebraska, illustrate the underlying daily exposure patterns (Figure 2C–D). In 2007, both counties had an annual mean of approximately 9.79 µg/m³, but their PMRI values were 28 and 16 and their P98 concentrations were 67.2 and 19.2 µg/m³, respectively. The calendar and ranked profiles show that similar annual means arose from markedly different daily exposure distributions. These county-years were selected solely from exposure data, without reference to mortality.

**Figure 2.**
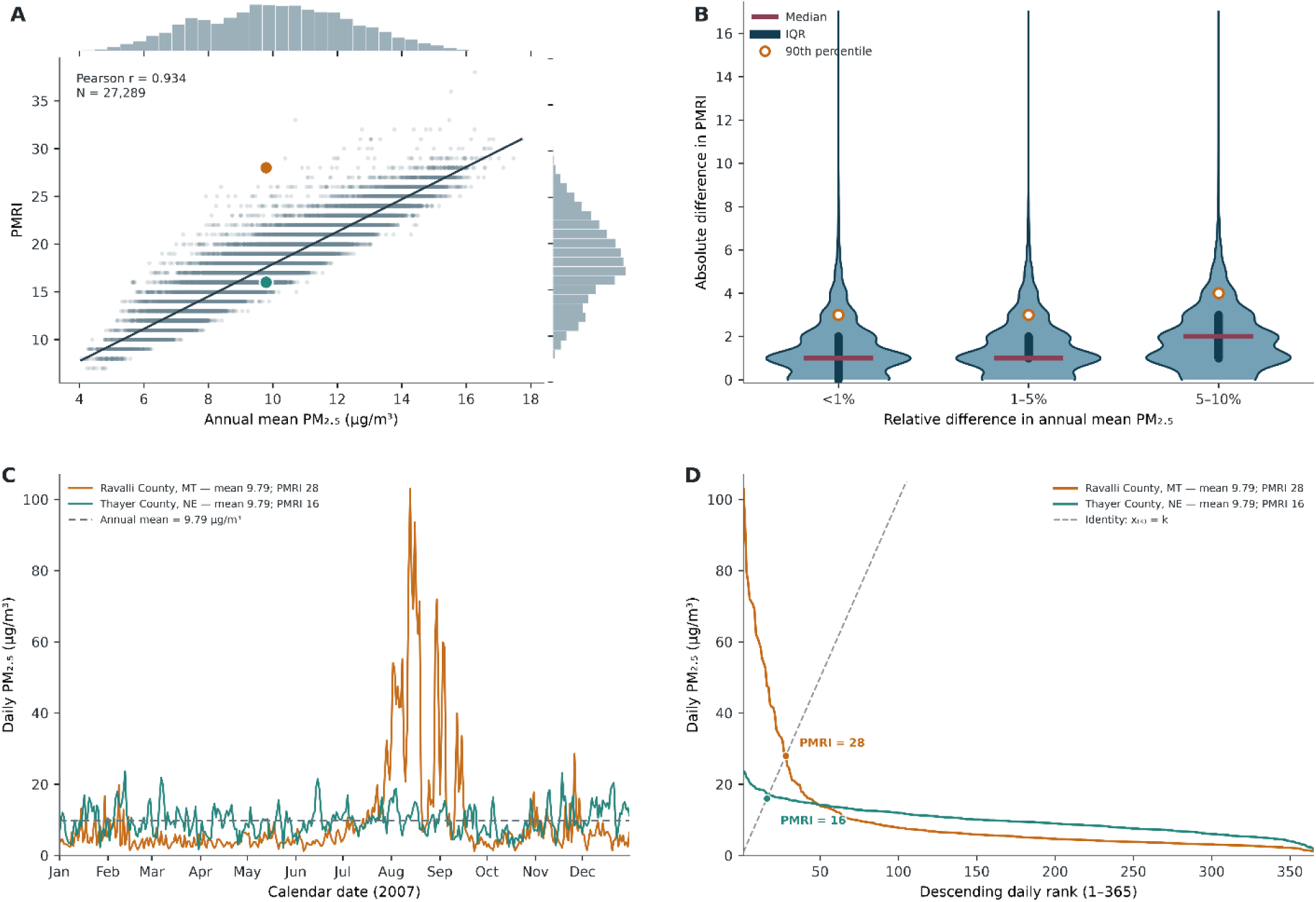
Annual mean PM_2.5_ and PMRI capture overlapping but non-equivalent features of the within-year exposure distribution. **A**, Relationship between annual mean PM_2.5_ and PMRI across 27,289 county-years. PMRI was strongly correlated with annual mean PM_2.5_ (Pearson r=0.934). Highlighted observations correspond to the two county-years shown in C and D. **B**, Distribution of absolute PMRI differences among same-year county-year pairs grouped by relative difference in annual mean PM_2.5_ (<1%, 1–5%, or 5–10%). Horizontal lines denote medians, vertical bars denote interquartile ranges, and open circles denote 90th percentiles. Pairwise distributions are descriptive because individual county-years can contribute to multiple pairs. **C**, Daily PM_2.5_ concentration profiles over the full 365-day calendar year for Ravalli County, Montana, and Thayer County, Nebraska, in 2007. Their annual mean PM_2.5_ concentrations were both approximately 9.79 µg/m³, whereas their PMRI values were 28 and 16, respectively; corresponding P98 concentrations were 67.2 and 19.2 µg/m³. The horizontal dashed line denotes the approximately common annual mean. **D**, The same 365 daily concentrations ranked from highest to lowest. The diagonal dashed identity line denotes x_(k)_=k; its intersection with each ranked concentration profile defines PMRI.

### 3.3 PMRI improves prediction beyond annual mean PM_2.5_

Although PMRI had lower standalone predictive performance than annual mean PM_2.5_ (held-out R², 0.0313 versus 0.0802), the two metrics performed substantially better in combination. In the primary county-grouped 10-fold analysis, adding PMRI to annual mean increased pooled held-out R² from 0.0802 to 0.1391, corresponding to an absolute ΔR² of 0.0589 (95% CI, 0.0500– 0.0677; Figure 3A; Table 2). This represented a 73.4% increase in R² relative to the annual-mean model.

**Figure 3.**
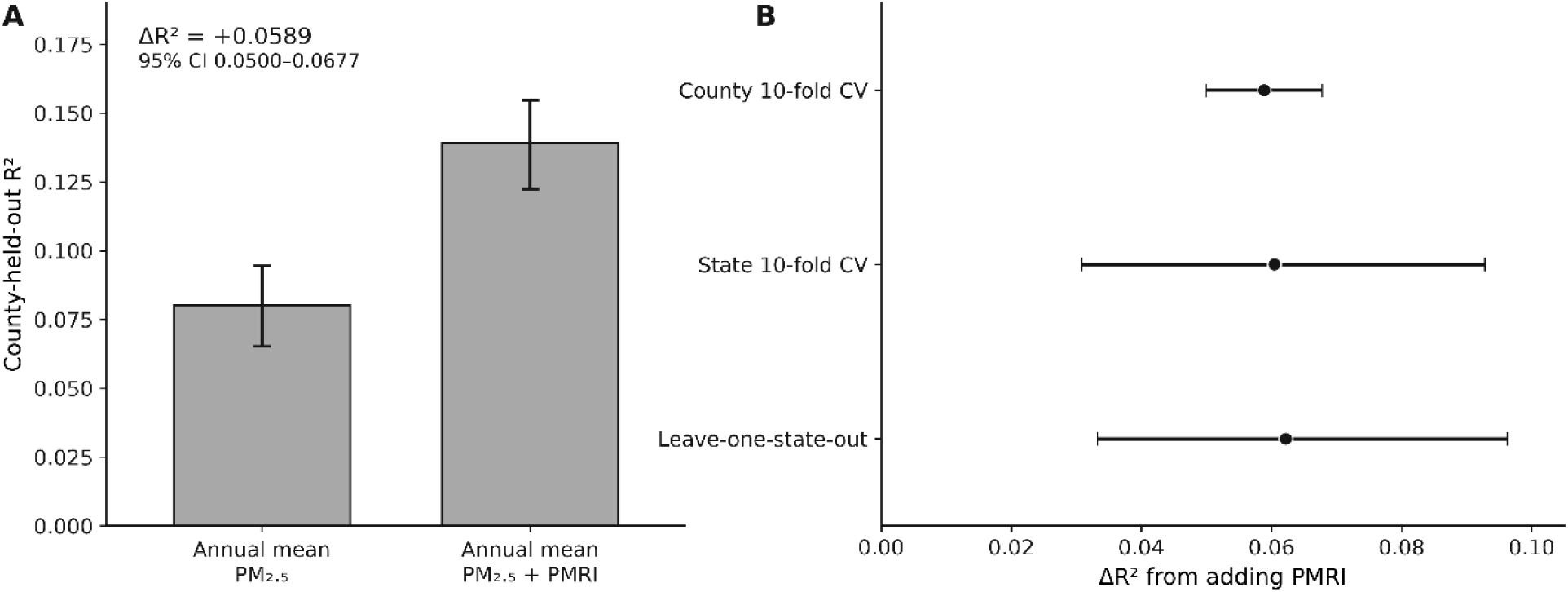
PMRI provides reproducible incremental mortality-predictive information beyond annual mean PM_2.5_. **A**, County-grouped held-out R² for annual mean PM_2.5_ alone and annual mean PM_2.5_ plus PMRI. Bars show pooled held-out R², and error bars indicate 95% confidence intervals obtained by geographic bootstrap resampling of counties. Adding PMRI increased held-out R² from 0.0802 to 0.1391, corresponding to a paired ΔR² of 0.0589 (95% CI, 0.0500–0.0677). **B**, Incremental held-out R² from adding PMRI under county-grouped 10-fold cross-validation, state-grouped 10-fold cross-validation, and leave-one-state-out validation. Points indicate pooled ΔR² estimates and horizontal lines indicate validation-appropriate geographic-bootstrap 95% confidence intervals. Positive increments were observed in all 10 county folds, 8 of 10 state folds, and 33 of 47 individually held-out states. ΔR² represents predictive-performance improvement and should not be interpreted as a mortality effect estimate.

**Table 2.** Geographic and temporal validation of PMRI.

| **Validation** | **Annual mean R<sup>2</sup>** | **Annual mean + PMRI R<sup>2</sup>** | **$\Delta R^2$ (95% CI)** | **$\Delta RMSE$** | **$\Delta MAE$** |
| --- | --- | --- | --- | --- | --- |
| County 10-fold CV | 0.0802 | 0.1391 | 0.0589 (0.0499–0.0677) | –4.74 | –4.70 |
| State 10-fold CV | 0.0683 | 0.1287 | 0.0604 (0.0308–0.0928) | –4.84 | –4.79 |
| Leave-one-state-out | 0.0614 | 0.1236 | 0.0622 (0.0332–0.0962) | –4.97 | –4.90 |
| Leave-one-year-out | 0.0771 | 0.1360 | 0.0589 (0.0386–0.0808) | –4.74 | –4.70 |
| 2003–2007 → 2008–2011 | 0.0556 | 0.1356 | 0.0800 (0.0701–0.0900) | –6.30 | –6.00 |
| 2003–2008 → 2009–2011 | 0.0505 | 0.1383 | 0.0878 (0.0773–0.0986) | –6.85 | –6.77 |

The base model included annual mean PM<sub>2.5</sub>, and the augmented model included annual mean PM<sub>2.5</sub> and PMRI. $\Delta R^2$ was calculated as $R^2_{\text{augmented}} - R^2_{\text{base}}$ . $\Delta RMSE$ and $\Delta MAE$ are augmented-minus-base differences; negative values favor the augmented model. Confidence intervals were obtained using paired cluster-bootstrap resampling of counties, states, or years, as appropriate. RMSE and MAE are expressed as deaths per 100,000 population. CI indicates confidence interval; CV, cross-validation; MAE, mean absolute error; RMSE, root mean squared error.

The improvement was consistent across performance measures and data partitions. Adding PMRI reduced RMSE by 4.74 deaths per 100,000 population and MAE by 4.70 deaths per 100,000 population. ΔR² was positive in all 10 county-held-out folds, indicating that the pooled improvement was not driven by a single partition. Thus, despite its weaker standalone performance, PMRI provided substantial predictive information conditional on annual mean PM_2.5_. The incremental performance was not explained by the functional form used for annual mean. When annual mean PM_2.5_ was represented by a training-fold-fitted three-knot cubic spline, adding PMRI yielded ΔR² = 0.0577 (95% CI, 0.0489–0.0667), similar to the linear-model result. PMRI also improved prediction after P98 was included with annual mean, although the increment was smaller (ΔR² = 0.0109; 95% CI, 0.0076–0.0142). These analyses indicate that the PMRI increment was not attributable to linear misspecification of annual mean and was not fully captured by a conventional upper-tail metric.

### 3.4 Geographic and temporal generalization

The incremental predictive performance of PMRI was retained when entire states were held out from model training (Figure 3B; Table 2). State-grouped 10-fold validation yielded ΔR² = 0.0604 (95% CI, 0.0308–0.0928), and pooled leave-one-state-out validation yielded ΔR² = 0.0622 (95% CI, 0.0332–0.0962). Both estimates were similar to the primary county-held-out increment of 0.0589, despite the greater geographic separation between training and evaluation data. Fold-and state-specific performance varied: ΔR² was positive in 8 of 10 state folds and 33 of 47 individually held-out states. Thus, improvement was not universal across individual states, but the positive pooled increments and confidence intervals excluding zero showed that the gain generalized beyond the counties and states used for model fitting.

Temporal analyses likewise showed that the incremental performance was retained across calendar periods. Pooled leave-one-year-out validation yielded ΔR² = 0.0589 (95% CI, 0.0386– 0.0808), nearly identical to the primary county-held-out estimate. Point estimates were positive for all nine held-out years, although the confidence interval for 2005 included zero. Forward validation produced somewhat larger increments: training on 2003–2007 and testing on 2008– 2011 yielded ΔR² = 0.0800 (95% CI, 0.0701–0.0900), whereas training on 2003–2008 and testing on 2009–2011 yielded ΔR² = 0.0878 (95% CI, 0.0773–0.0986). The corresponding reductions in RMSE were 6.30 and 6.85 deaths per 100,000 population. Because both forward analyses evaluated models exclusively in subsequent years, these findings provide direct evidence that the incremental performance of PMRI was not confined to contemporaneous or randomly partitioned data.

### 3.5 Predictive performance across mortality outcomes

The incremental predictive performance of PMRI was not limited to all-cause mortality. In county-grouped validation, adding PMRI improved held-out prediction for each of the six prespecified cause-specific outcomes, with positive ΔR² estimates and bootstrap confidence intervals excluding zero throughout (Figure 4; Table 3). The all-cause increment was 0.0589 (95% CI, 0.0500–0.0677), and cause-specific increments ranged from 0.0159 to 0.0600. For cardiovascular or circulatory mortality, ΔR² was 0.0424 (95% CI, 0.0352–0.0494). Within this category, the increments were 0.0159 (0.0100–0.0215) for ischemic heart disease, 0.0508 (0.0395–0.0620) for acute myocardial infarction, and 0.0531 (0.0415–0.0649) for stroke. PMRI also improved prediction of respiratory mortality, with ΔR² = 0.0600 (0.0482–0.0715), and of the more specific chronic lower respiratory mortality outcome, with ΔR² = 0.0597 (0.0441–0.0748). Thus, the improvement was evident for broad cardiovascular and respiratory groupings as well as for more narrowly defined causes of death. The magnitude of the increment varied across outcomes. Ischemic heart disease had the smallest county-held-out gain, whereas respiratory mortality had the largest. These values are not directly comparable as measures of etiologic importance because the outcome-specific cohorts differed in size, mortality-rate variability, and baseline predictive performance.

**Figure 4.**
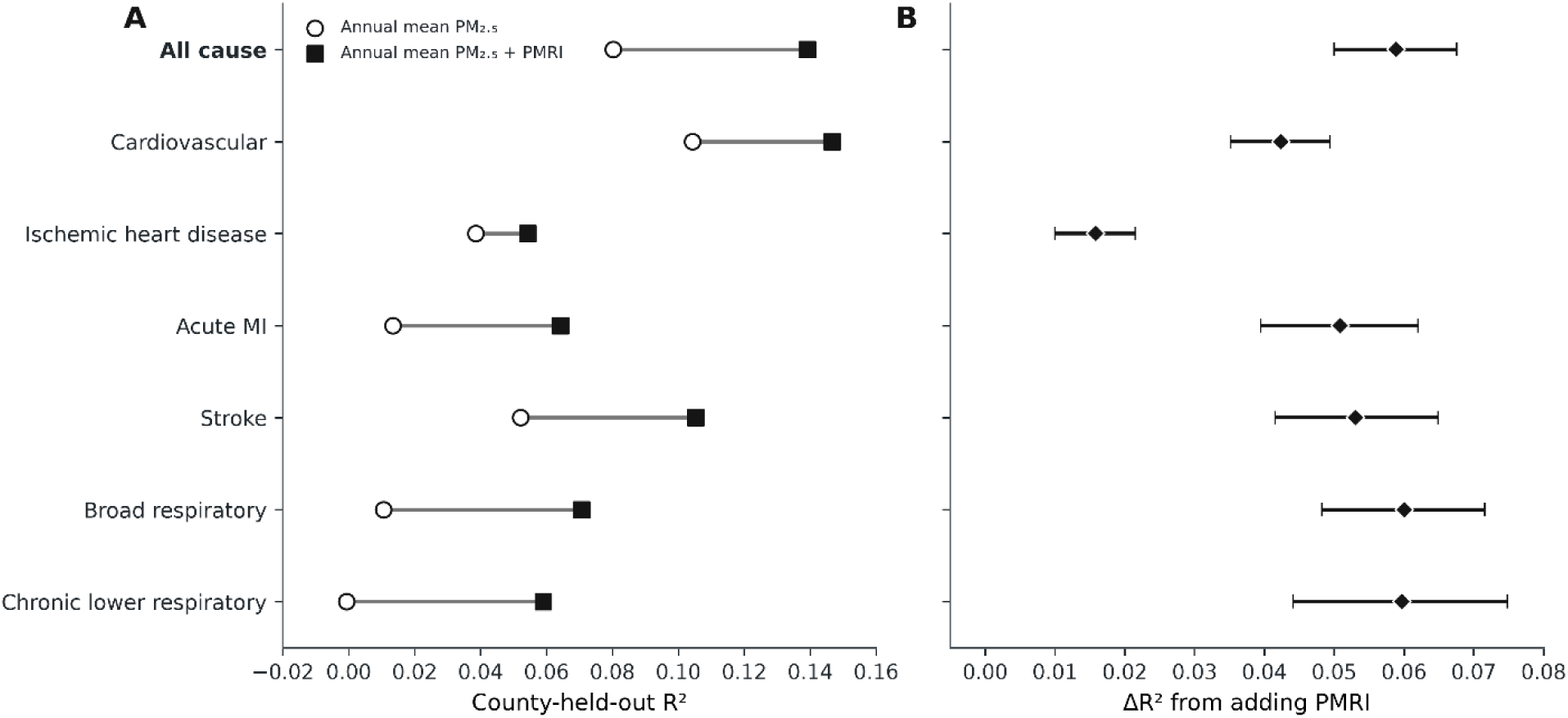
PMRI improves geographically held-out prediction across mortality outcomes. **A**, County-grouped held-out R² for annual mean PM_2.5_ alone and annual mean PM_2.5_ plus PMRI for all-cause mortality and six cause-specific mortality outcomes. Open circles indicate annual mean PM_2.5_ alone, filled squares indicate annual mean PM_2.5_ plus PMRI, and connecting lines link the paired model performances within each outcome. **B**, Corresponding incremental held-out R² from adding PMRI. Diamonds indicate paired ΔR² estimates and horizontal lines show 95% confidence intervals obtained by geographic bootstrap resampling of counties. All-cause mortality was the primary endpoint; cause-specific analyses assess generalization of the predictive increment. ΔR² is a predictive-performance measure and should not be interpreted as a mortality effect estimate.

**Table 3.** County-held-out predictive performance across mortality outcomes.

| **Outcome** | **County-**<br>**years** | **Annual**<br>**mean R<sup>2</sup>** | **Annual mean +**<br>**PMRI R<sup>2</sup>** | **$\Delta R^2$ (95% CI)** |
| --- | --- | --- | --- | --- |
| All-cause mortality | 27,289 | 0.0802 | 0.1391 | 0.0589 (0.0499–0.0677) |
| Cardiovascular or circulatory mortality | 24,970 | 0.1043 | 0.1467 | 0.0424 (0.0352–0.0494) |
| Ischemic heart disease | 20,877 | 0.0385 | 0.0544 | 0.0159 (0.0100–0.0215) |
| Acute myocardial infarction | 13,140 | 0.0133 | 0.0642 | 0.0508 (0.0395–0.0620) |
| Stroke or cerebrovascular mortality | 11,843 | 0.0522 | 0.1053 | 0.0531 (0.0415–0.0649) |
| Respiratory mortality | 17,114 | 0.0106 | 0.0707 | 0.0600 (0.0482–0.0715) |
| Chronic lower respiratory mortality | 11,884 | –0.0006 | 0.0591 | 0.0597 (0.0441–0.0748) |

Each outcome was analyzed in its own eligible cohort after exclusion of suppressed, missing, or unreliable mortality rates. Models compared annual mean PM<sub>2.5</sub> alone with annual mean PM<sub>2.5</sub> plus PMRI. Confidence intervals for $\Delta R^2$ were obtained by paired cluster-bootstrap resampling of counties. $\Delta R^2$ was calculated from unrounded R<sup>2</sup> estimates; therefore, differences between displayed rounded values may not exactly equal the reported $\Delta R^2$ . All-cause mortality was the primary outcome. $\Delta R^2$ is a predictive-performance measure and should not be interpreted as a mortality effect estimate. CI indicates confidence interval; PMRI, PM<sub>2.5</sub> magnitude-rank index.

Geographic validation of the cause-specific findings produced a similar overall pattern. In secondary state-grouped analyses, ΔR² point estimates were positive for all seven outcomes, and six of the seven state-bootstrap confidence intervals excluded zero. Ischemic heart disease was the exception: its state-held-out estimate remained positive but was less precise (ΔR² = 0.0129; 95% CI, −0.0099 to 0.0336). Collectively, these results show that the incremental performance of PMRI extended across multiple causes of death, although the strength and geographic precision of the evidence varied by outcome.

### 3.6 Additional sensitivity analyses

Two additional analyses examined whether the primary result was sensitive to the influence of highly populous counties or to the integer definition of PMRI. Excluding the 1% largest counties by population yielded ΔR² = 0.0576, compared with 0.0589 in the primary analysis. Sequential omission of each of the 10 largest counties produced similar results and did not identify any single county as the source of the incremental performance. Using the continuously interpolated PMRI defined in the Methods produced a modest additional improvement. Held-out R² increased from 0.13910 for integer PMRI to 0.14172 for continuous PMRI, corresponding to ΔR² = 0.00262 (95% CI, 0.00149–0.00369). RMSE decreased by a further 0.215 deaths per 100,000 population and MAE by 0.183. Although the confidence interval excluded zero, this gain was only approximately 4.4% of the original increment obtained by adding integer PMRI to annual mean and did not materially alter the primary conclusion.

## 4. Discussion

Annual mean PM_2.5_ was the strongest standalone predictor among the prespecified exposure metrics, but it did not capture all mortality-predictive information in the daily exposure distribution. Adding the outcome-independent PMRI increased county-held-out R² from 0.0802 to 0.1391 (ΔR² = 0.0589; 95% CI, 0.0499–0.0677) and reduced both RMSE and MAE. Similar increments were observed in state-held-out, year-held-out, and forward-time validation, and positive county-held-out increments were found for all-cause mortality and all six cause-specific outcomes. The persistence of the improvement across locations, periods, and outcomes indicates that within-year magnitude–frequency structure contained predictive information beyond annual mean concentration.

Long-term mean PM_2.5_ exposure is consistently associated with mortality [1–6], and annual mean was likewise the strongest standalone predictor in the present analysis. However, aggregation across days obscures within-year variation: similar annual means can arise from persistent moderate concentrations or from lower background concentrations interrupted by high-concentration episodes. These profiles differ in the magnitude and frequency of elevated daily exposure. Studies spanning acute and chronic timescales, together with analyses of exposure variability, extreme pollution days, and wildfire-related pollution, indicate that such distributional features may be relevant to mortality [9–17]. The PMRI increment may capture the combined influence of emission sources, seasonality, meteorology, exposure measurement, behavioral responses, and population susceptibility. More broadly, the results indicate that reducing daily exposure data to an annual mean can discard information relevant to mortality prediction and support evaluating distribution-aware summaries alongside annual mean exposure.

The consistency of the PMRI increment across validation designs supports its generalizability. County grouping prevented different years from the same county from entering both training and evaluation data, while state-held-out and temporal analyses tested transfer across larger regions and calendar periods. Similar pooled ΔR² estimates and positive increments across all examined mortality outcomes indicate that the improvement was not confined to particular counties, states, years, or outcomes. However, improvement was not observed in every held-out state, and the state-level confidence interval for ischemic heart disease included zero; generalization was therefore broad but not uniform. PMRI was prespecified and defined independently of mortality rather than optimized for prediction. Its smaller increment after inclusion of P98 indicates partial overlap with an upper-tail metric, while the modest benefit of continuous interpolation supports retention of the simpler integer definition. PMRI should therefore be considered an informative complement to annual mean exposure in this setting, not a uniquely optimal metric.

Strengths of this study include nationwide coverage, an outcome-independent PMRI definition, a strong annual-mean benchmark, and geographic and temporal held-out validation. The ecological design, modeled county-level exposures, and absence of confounder adjustment preclude individual-level or causal interpretation; the PMRI increment may partly reflect spatial patterning or correlated determinants of mortality. Mortality suppression reduced some cause-specific cohorts, the 2003–2011 period may not represent current pollution patterns, and performance varied across states and years. Future studies should evaluate prespecified distribution-aware metrics in individual-level cohorts, more recent periods, and geographically independent populations with explicit confounder adjustment.

## 5. Conclusion

Annual mean PM_2.5_ was a strong standalone predictor but did not capture all mortality-predictive information in the within-year distribution of daily exposure. Adding PMRI improved out-of-sample prediction across held-out counties, states, and years and across mortality outcomes.

These findings support evaluating within-year exposure structure alongside annual mean concentration in studies of long-term PM_2.5_ exposure and mortality.

## Statements and Declarations

### Ethical approval

Not applicable.

### Consent to Participate

Not applicable.

### Author Contributions

R.Z. conceptualized, designed the study, analyzed the data, and wrote the paper.

### Ethical Responsibilities of Authors

The author has read, understood, and complied, as applicable, with the statement on Ethical Responsibilities of Authors in the Instructions for Authors.

### Data Availability Statement

Daily county-level PM_2.5_ concentrations are publicly available from the CDC Environmental Public Health Tracking Program (https://data.cdc.gov/d/53mz-4zqd). County-level mortality data are publicly available from the CDC WONDER Underlying Cause of Death database (https://wonder.cdc.gov/ucd-icd10.html).

